# A Wearable Plantar Pressure System for Early Warning of Freezing of Gait Based on Time-Frequency and State-Space Modeling

**DOI:** 10.64898/2026.06.30.26356907

**Authors:** Yifan Yuan, Wenhao Li, Liujinxiang Zhu, Hechong Su, Hailong Yu, Han Wang, Guan Ning Lin

## Abstract

Freezing of gait (FoG) in Parkinson’s disease is a brief but hazardous gait failure that often precedes falls. For wearable cueing or other closed-loop assistance, a detector that reacts only after FoG onset is usually too late; the more useful task is to recognize the pre-freezing transition from physiological signals. This study presents PreFoGNet, a dual time-frequency deep learning framework for early FoG prediction using plantar pressure signals. The temporal stream combines a multi-scale Inception encoder with a bidirectional Mamba module to capture both short contact-related transients and several-second gait deterioration without the quadratic cost of attention. In parallel, the frequency stream uses band-wise spectral modeling and attention-based gating to emphasize physiologically meaningful changes in the locomotion, freeze-related, and high-frequency bands. On the WearGait-PD dataset, with a 2 s prediction horizon and subject-wise evaluation, PreFoGNet achieved a sensitivity of 93.94%, a specificity of 89.76%, a G-Mean of 0.9183, and an AUC-ROC of 0.9607. It outperformed classical machine-learning and deep learning baselines, and retained usable performance under moderate noise and single-channel loss. Additional horizon analysis showed that plantar pressure contains a stable pre-freezing signature within 0-3 s before onset, with a practical prediction boundary of approximately 6-7 s. These findings suggest that time-frequency modeling of plantar pressure is a promising signal-processing route for wearable FoG early-warning systems.

## I. Introduction

PARKINSON’S disease (PD) is a progressive neurodegenerative disorder in which gait impairment increasingly limits independent mobility and quality of life [1, 2]. Among its motor complications, freezing of gait (FoG) is particularly disabling. FoG is commonly described as a sudden and brief inability to generate effective forward stepping despite the intention to walk [3, 4], and it is closely associated with falls, fear of falling, and reduced community participation [5]. Pharmacological treatment does not fully control FoG in many patients, especially in advanced disease. For this reason, wearable monitoring and cueing systems have attracted growing interest [6]. However, most systems detect FoG after the freezing episode has already started. At that point, the opportunity to deliver a preventive cue may have been missed. The key signal-processing problem is therefore not only FoG detection, but the prediction of an impending transition from regular gait to freezing.

Plantar pressure is a useful modality for this purpose because it measures the mechanical interaction between the foot and the ground. Unlike EEG, it is less affected by neural recording artifacts; unlike IMU-based measurements, it is less dependent on sensor orientation and placement on the body. Pressure insoles also capture bilateral load transfer, heel strike, midfoot loading, and toe-off dynamics, all of which may change before a freezing episode. The challenge is that these changes are subtle, transient, and embedded in high-dimensional non-stationary signals. A model designed for this task should therefore extract predictive information from both the temporal evolution of gait and the spectral structure of plantar pressure fluctuations.

Previous work has made important progress but leaves two methodological gaps. First, early prediction requires a model that can preserve weak prodromal information over several seconds. CNN-based methods are effective at extracting local waveform patterns, but their receptive fields are often too limited to represent the gradual deterioration of gait rhythm over longer horizons [7]. LSTM-based models address sequential dependence, yet their recurrent computation and memory decay can make it difficult to retain faint early cues in high-frequency, multi-channel pressure sequences [8]. Transformer models offer global context, but their computational burden and data demand may be problematic for imbalanced FoG datasets and future wearable deployment.

Second, many end-to-end models treat the pressure sequence as a generic time series and do not explicitly use physiological knowledge about FoG [9]. This is important because pre-freezing must be separated from turning, hesitation, shuffling, or other non-freezing gait irregularities [10]. A high false-positive rate is not a minor issue in a wearable system: repeated unnecessary cues may reduce adherence and trust. Prior studies have linked FoG to abnormal rhythmic fluctuations and energy redistribution in specific frequency ranges [11]. Incorporating this prior information may therefore improve specificity while keeping the high sensitivity required for safety-critical monitoring.

A further open question is the useful prediction horizon. Reporting performance at a single lead time does not tell us when a cue should be triggered or how prediction quality decays as the system looks further ahead. Although plantar pressure has been shown to support short-term FoG prediction [12], the effective boundary of this signal has not been systematically quantified. This boundary is clinically relevant because different lead times may support different intervention strategies, from mild prompts at an early stage to stronger rhythmic cueing closer to the freezing onset [13].

To address these issues, we propose PreFoGNet, a dual-stream framework tailored to plantar-pressure-based FoG prediction. The frequency stream first examines whether PreFoG has distinct spectral signatures and then models the spectrum through physiologically defined bands with an attention-based gating mechanism. The temporal stream combines multi-scale convolutional encoding with a bidirectional Mamba module to represent both local contact events and longer gait deterioration. We evaluate the method using subject-wise data splitting, compare it with classical and deep learning baselines, analyze prediction horizons and window lengths, and test robustness to noise and sensor loss. The aim is to provide not only a predictive model, but also a clearer signal-processing account of when and where plantar pressure carries information about upcoming FoG.

### II. Materials and methods

### A. Problem definition

We defined early FoG prediction as a subject-wise binary classification problem using multichannel plantar-pressure windows. The task is a forecasting task rather than a detection task: the model uses the pressure history observed up to the current time and estimates whether a FoG onset will occur after the window ends.

Let 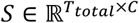 denote the collected raw plantar pressure signals, where *T*_*total*_is the total number of sampling points and *C* is the number of sensor channels (*C =* 32, corresponding to the plantar pressure sensor arrays). We employ a non-overlapping sliding window technique to segment the continuous signal. An input sample *X*_*t*_ ∈ ℝ^*L*×*C*^ represents an observation window of length *L* (e.g., *L =* 100, corresponding to 1*s* of data) ending at the current time *t*, capturing historical gait dynamics from time *t* − *L* + 1 to *t*.

Let 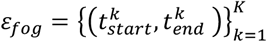 denote the start and end times of *K* clinical expert-annotated FoG events. Unlike traditional FoG detection, which classifies whether the current window itself is a freezing episode, our objective is early forecasting: given the observation window *X*_*t*_, the goal is to predict whether a FoG event will onset within a future “prediction horizon” (denoted as *T*_*horizon*_). Consequently, the predictive label *y*_*t*_ ∈ {0,1} for the input window *X*_*t*_ is defined as follows:

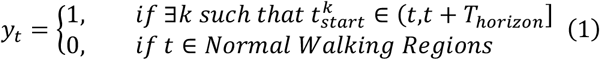

Here, *y*_*t*_ *=* 1 indicates the PreFoG class, meaning a freezing event will occur within the next *T*_*horizon*_ seconds (e.g., 2*s*); *y*_*t*_ *=* 0 denotes the normal walking class, which includes normal walking, turning, or non-freezing gait abnormalities (such as staggering). To ensure the absolute purity of negative samples and avoid label confusion, we introduce a safety margin to guarantee that the designated normal walking regions are strictly isolated from any pre-freezing or freezing periods.

### B. Dataset and preprocessing

#### 1) Dataset Description

The data for this study are derived from the public dataset WearGait-PD [14]. This dataset encompasses gait data from 100 Parkinson’s disease (PD) patients and 85 age-matched controls. All subjects have medication usage and clinical assessment information from medical visit reports or self-reports, and completed standardized gait tests according to the protocols and requirements.

The core equipment for data acquisition is the Moticon OpenGo smart sensor insole. Each insole integrates 16 distributed pressure sensors capable of capturing plantar pressure distribution at a sampling rate of 100 Hz. The clinical experts performed frame-by-frame annotations of gait behaviors through synchronized video recordings to ensure data synchronization and alignment.

To encompass diverse gait patterns and include as many freezing of gait events as possible, we selected six walking tasks from the dataset closely related to daily life, including: Self-Paced Walking, Hurried-Paced Walking, Walking with Turns, and the Timed-Up-and-Go (TUG) test. The above task designs can effectively capture the kinetic characteristics of PD patients during straight walking, turning, and gait initiation.

#### 2) Data Preprocessing

Preprocessing was kept close to the measurement process. Missing values caused by wireless packet loss or brief poor sensor contact were filled by linear interpolation along each channel. Inactive segments at the beginning and end of each trial were then removed using the sum of absolute changes across pressure channels. To reduce between-subject differences in body size, pressure values were normalized by body weight and treated as relative load distributions.

To investigate the impact of different input time windows on the capability to capture pre-freezing signs and to determine the optimal input sequence length, we construct a multi-scale time series dataset. We employ the sliding window technique to segment the continuous pressure signal stream into time fragments *X* ∈ ℝ^*L*×*C*^. This study constructs three independent data subsets corresponding to specific window lengths *L* ∈ {100,200,300} (representing data sampling durations of 1*s*, 2*s*, and 3*s*, respectively), aiming to analyze the trade-off in the model’s ability to capture short-term local sudden changes versus long-term progressive evolutionary features.

Based on clinically annotated events, we formulated the following sample generation logic: for continuous segments labeled as “FoG”, samples are extracted using non-overlapping segmentation and marked as the freezing period (FoG); To achieve fine-grained early prediction, we set a lookback horizon of *T*_*horizon*_ *=* 10*s*. A sliding window sampling with a step size of *S =* 100 (i.e., 1*s*) is employed to generate a series of samples with different lead times. For any FoG event onset *t*_*start*_, we extract the data within the interval [*t*_*start*_ − *L* − (*i* · *S*), *t*_*start*_ − (*i* · *S*)] as samples labeled as *ProFog*_*i*_, where *i* ∈ {0,1, …,9}. When *L* > *S* (e.g., *L =* 200,300), adjacent PreFoG samples partially overlap in the time domain); To construct high-confidence negative samples, we implement a dynamic safety boundary strategy, introducing a safety buffer period; a segment of normal walking data is included as a negative sample only when its distance from any expert-annotated freezing onset and freezing offset exceeds 10 seconds. This strategy effectively eliminates the potential prodromal phase and post-freezing recovery phase, ensuring the purity of normal samples and avoiding the misleading effects of label noise on model training.

Ultimately, each sample *X* ∈ ℝ^100×32^ contains 32 channels of pressure features (16 sensors on each foot).

### C. Overview of PreFoGNet

PreFoGNet was designed around a practical signal-processing assumption: the pre-freezing transition is expressed both in how pressure evolves over time and in how its energy is distributed across frequency bands. As shown in Fig. 1, each window is processed by two branches. The temporal branch extracts contact-related morphology and gait evolution, whereas the frequency branch extracts band-wise spectral information. The two outputs are fused for binary PreFoG prediction.

**Fig. 1.**
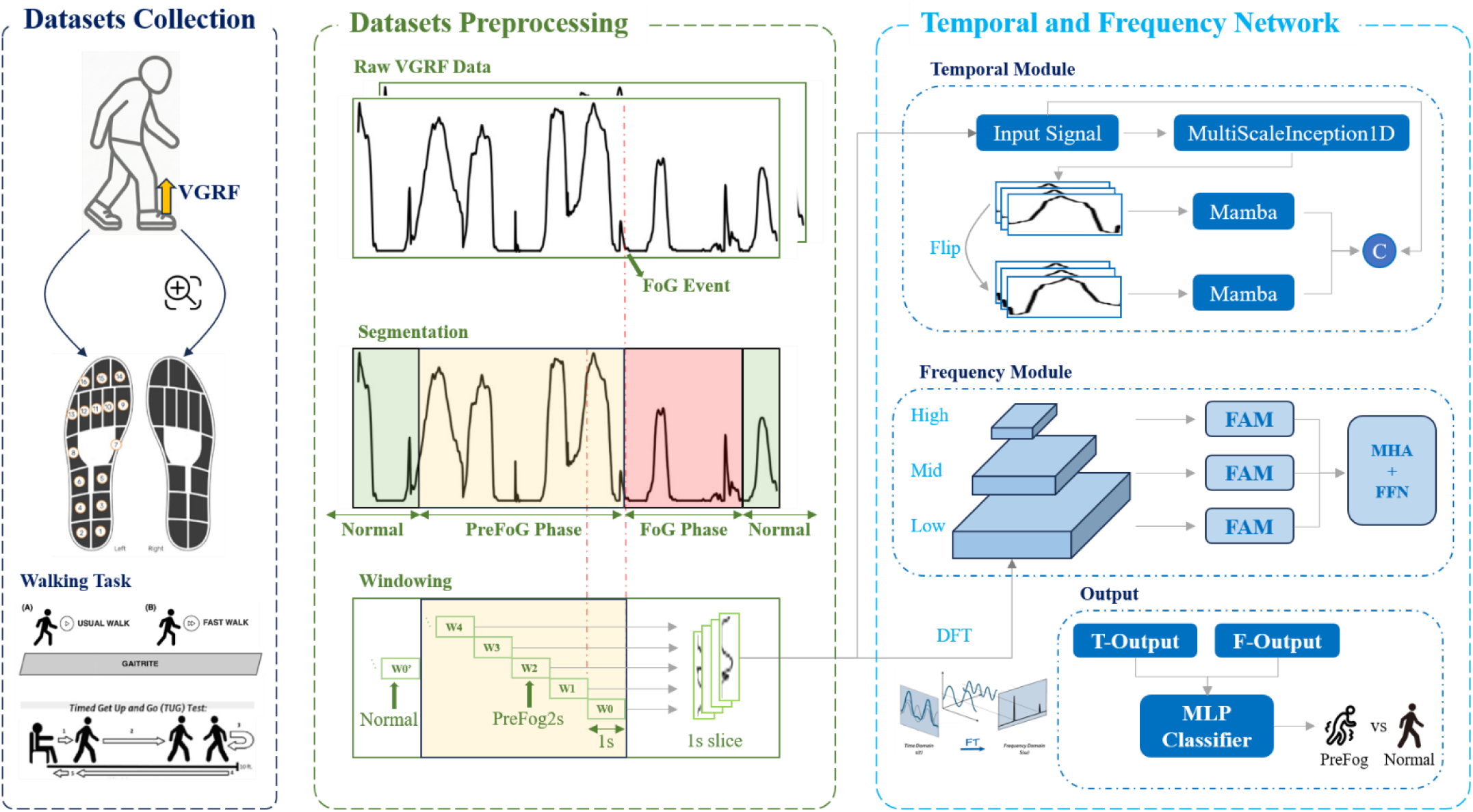
Overview of PreFoGNet

In the temporal branch, multi-scale one-dimensional convolutions first describe pressure patterns at several durations, from brief contact transients to broader gait-cycle changes. A bidirectional Mamba block then summarizes the encoded sequence over the full observation window. In the frequency branch, the window is transformed by DFT and divided into gait-related bands: low-frequency locomotor rhythm, the 3-8 Hz freeze-related range, and higher-frequency components.

The final classifier receives three sources of information: the long-range temporal representation, a shallow residual temporal representation, and the band-wise spectral representation. This structure keeps the model close to the signal: improvements or errors can be discussed in terms of temporal context, local pressure morphology, or spectral content rather than as an undifferentiated latent embedding.

### D. Gait Dynamics Modeling via Temporal Stream

Plantar-pressure contain events at different time scales. Heel strike and toe-off occur over short intervals, while changes in rhythm and load transfer may unfold over several steps. As shown in Fig. 2, the temporal stream was therefore built to preserve both local contact details and longer pressure history.

**Fig. 2.**
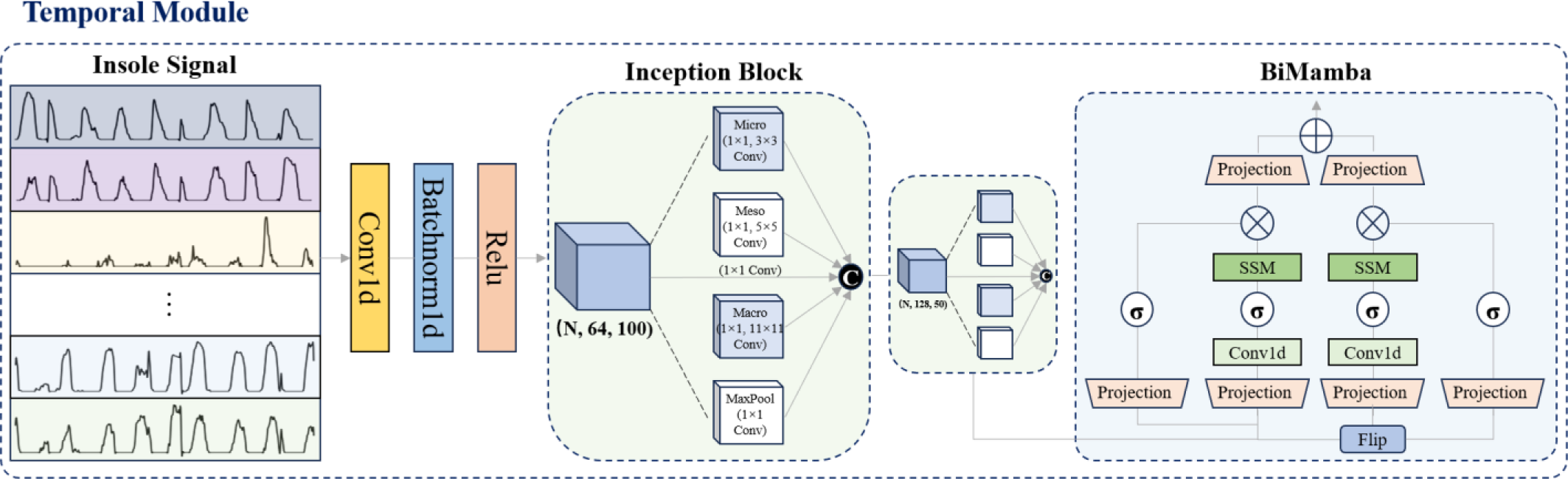
Temporal stream of PreFoGNet

To decouple the mixed features of transient ground contact impacts (high frequency) and gait cycle rhythms (low frequency) in gait signals at the physical level, we constructed a multi-scale Inception encoder ℱ_*inc*_ [15]. This design simulates the diagnostic paradigm of clinical experts, namely paying simultaneous attention to micro-motor abnormalities and macro-gait dynamics.

Let the *c* channel of the input signal be *x*^(*c*)^; this encoder contains *K =* 4 parallel feature extraction branches. For the *k* − *th* branch, we employ 1D convolution operations using convolution kernels 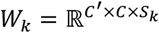 of different sizes. The output feature map *Z*_*k*_ of the *k* − *th* branch is defined as:

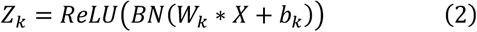

where * denotes the convolution operator, and *BN*(·) is batch normalization. The design of the convolution kernel size *S*_*k*_, which directly corresponds to different pathological receptive fields: Micro-Scale (*S*_*k*_ *=* 3) captures high-frequency transient ground contact mutations and early tremor signals; Meso-Scale (*S*_*k*_ *=* 5) captures local transitions in gait phases (from stance to swing phase); Macro-Scale (*S*_*k*_ *=* 11) captures the complete gait cycle envelope; the Pooling Branch utilizes a max-pooling layer to extract salient features and suppress sensor noise. The final local feature *H*_*local*_ is obtained through concatenation along the channel dimension, thereby forming a comprehensive representation of multi-scale gait dynamics.

After acquiring the local features, it is necessary to establish long-range dependencies spanning a long window (e.g., 300 steps) to identify the early progressive signs of FoG. To this end, we introduce Bi-Mamba, utilizing the Selective State Space Model [16] to capture slowly varying gait deterioration trends while preserving computational efficiency. The core of Mamba’s modeling of gait evolution lies in its dynamic process of mapping the one-dimensional input signal *x*(*t*) to the output *y*(*t*), and accomplishing the characterization of gait temporal features through the sequential transmission and update of the latent state *h*(*t*):

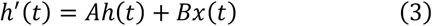

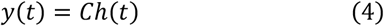

where *A* ∈ ℝ^N×N^ is the state transition matrix, determining the system’s memory decay rate for historical gait, and *B* ∈ ℝ^N×1^, *C* ∈ ℝ^1×N^ are the projection parameters for the input and output, respectively.

To process the discretely sampled sensor data, Mamba utilizes the zero-order hold (ZOH) principle and introduces a time scale parameter Δ to discretize the above equations. Unlike traditional LTI systems, Mamba employs a Selection Mechanism, meaning that the parameters *B, C*, Δ are no longer static but act as functions of the input *H*_*local*_ :

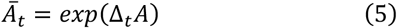

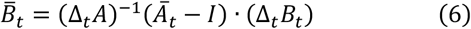

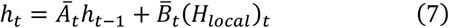

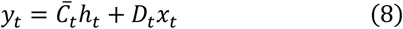

Here, Ā_*t*_ and 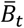 represent the discretized state transition matrix and input matrix, Δ_*t*_∈ ℝ^D^ is the time scale parameter, *I* is the identity matrix, and *h*_*t*_ ∈ ℝ^D×N^ is the latent state at the current time step, which compresses and stores the complete gait dynamics context from time 0 to *t*; 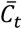 is the input-dependent output projection matrix responsible for the selective reading of the accumulated historical state *h*_*t*_ to extract long-range temporal features, while *D*_*t*_ constitutes a gated residual connection used to preserve the instantaneous information in the original input *x*_*t*_. This input-dependence endows the model with the capability of selective state updating, allowing it to adaptively regulate the information flow based on current gait dynamic features, thereby effectively suppressing irrelevant walking artifacts while preserving crucial pre-freezing signs. We therefore used a bidirectional structure: one state-space block processes the encoded sequence in chronological order, and the other processes the reversed sequence:

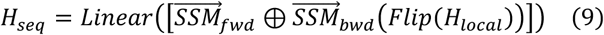

where *Flip*(·) denotes the time-reversal operation along the temporal dimension. To obtain a fixed-length compact representation as input for the classifier, we apply Global Average Pooling (GAP) to the sequential features, aggregating them into a compact temporal context vector *v*_*temporal*_; this process achieves a linear computational complexity of *O*(*L*) and maximizes the effective receptive field while maintaining computational efficiency for wearable deployment.

### E. Spectral modelling in the frequency stream

The frequency stream was included to make use of rhythm and energy changes that may be difficult to capture from the time waveform alone. As shown in Fig. 3, its processing has three steps: transformation to the frequency domain, separation into gait-related bands, and band-level reweighting.

**Fig. 3.**
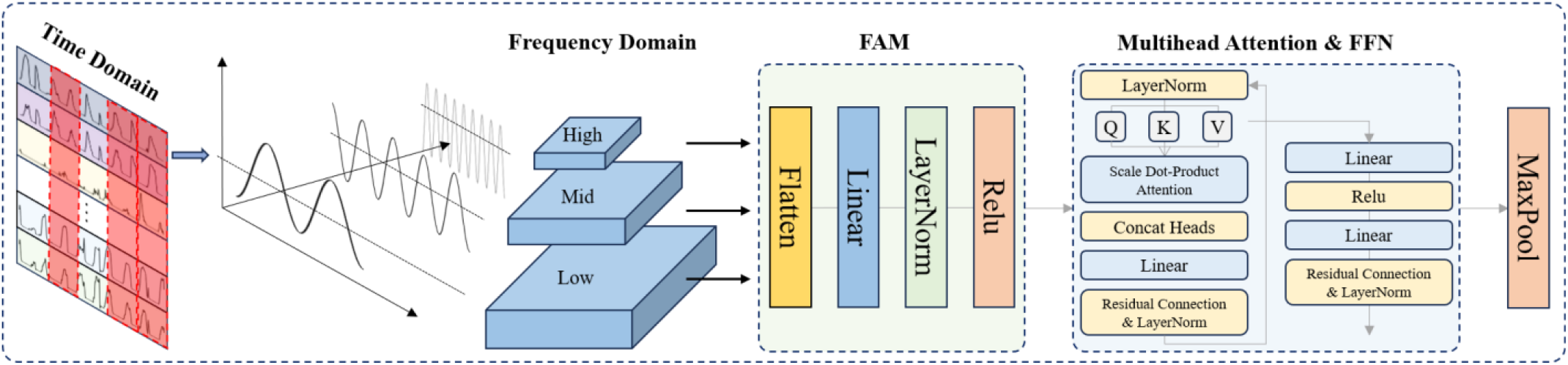
Frequency stream of PreFoGNet

First, to obtain the global frequency distribution, we apply the Discrete Fourier Transform (DFT) to the input plantar pressure signal *X* ∈ ℝ^*L*×*C*^. For the sequence *x*^(*c*)^[*n*] of the *c* − *th* channel, its spectral representation *F*^(*c*)^[*k*] is calculated as follows:

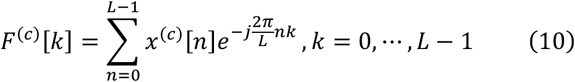

To map the discrete index *k* to the physical frequency *f*_*k*_, we define 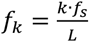, where *f*_s_ is the sampling rate (e.g., 100 Hz). To enhance numerical stability and emphasize weak harmonic components, we calculate the logarithmic amplitude spectrum, introducing a small constant *ϵ* to prevent logarithmic divergence:

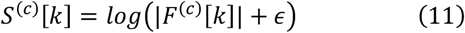

nsidering the physical characteristics of human plantar movement, we implement a hard-threshold low-pass filter in the frequency domain, truncating high-frequency components with *f* > 20*Hz* to remove environmental and sensor thermal noise. Rather than treating the entire filtered spectrum *S*_*filtered*_ as an undifferentiated feature vector, we explicitly divide it into three sub-bands based on established clinical priors regarding Parkinsonian gait [17]. The Locomotion Band (*B*_*low*_, 0-3 Hz) encodes the basic stepping cadence and gait cycle during normal walking. The Pathological Freeze Band (*B*_*mid*_, 3-8 Hz) serves as the critical region for FoG prediction, as energy surges in this band physiologically correspond to high-frequency alternating tremors when effective forward stepping is blocked. The High Band (*B*_*high*_, 8-20 Hz) contains rapid postural adjustment harmonics and non-freezing abnormal artifacts.

To accommodate the inconsistent bandwidths, we adopt a decoupled design: each sub-band *B*_*i*_ (*i* ∈ {*low, mid, high*}) is mapped into a unified embedding space through an independent linear projection layer *ϕ*_*i*_, generating the band embedding vector *E*_*i*_ *= ϕ*_*i*_(*B*_*i*_). This allows the model to independently learn band-specific physiological patterns.

During the PreFoG stage, pathological signs manifest not only as localized energy variations but also as complex spectral energy redistributions. To capture these correlations, we stack the three band embedding vectors into a sequence *E =* [*E*_*low*_, *E*_*mid*_, *E*_*high*_]*T* and introduce a spectral gating mechanism based on Multi-Head Self-Attention [18]. In this formulation, attention is computed across the frequency bands rather than spatial channels. The query (*Q*), key (*K*), and value (*V*) matrices are generated via linear transformations of *E*, and the gated output is computed as:

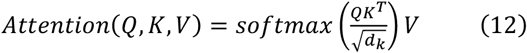

This design gives the network a simple way to combine spectral bands according to the pattern present in the current window.

### F. Multi-Dimensional Feature Fusion and Classification

The two streams were fused after branch-specific feature extraction. This delayed fusion allows the temporal stream to summarize pressure evolution and the frequency stream to summarize spectral content before the classifier combines them. We concatenate the three key feature vectors from the two-stream architecture to construct a high-dimensional mixed feature vector *V*_*fusion*_. Herein, the global sequence context (*v*_*seq*_) is derived from the output of the Bi-Mamba module, containing gait evolution trends and temporal feature information spanning several seconds; the local residual feature (*v*_*res*_) originates from the skip connection of the shallow layer in the temporal stream (Multi-Scale Inception output), and to align with the global feature dimension, we apply Global Average Pooling (GAP) after the Inception output. The spectral feature (*v*_*freq*_) comes from the output of the frequency stream, encompassing inter-band interactive representations generated by the multi-head attention mechanism to capture nonlinear correlations among sub-bands.

To mine the high-order nonlinear interactive relationships among features, the concatenated vector *V*_*fusion*_ is fed into a Multilayer Perceptron (MLP) classifier. This classifier contains two hidden layers with 256 and 128 nodes, respectively. We introduce Batch Normalization (BN) and Dropout mechanisms after each linear layer.

The objective of PreFoGNet is to learn a nonlinear mapping function ℱ: ℝ^*L*×*C*^ → [0,1]. Given a dataset 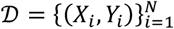 containing N samples, we need to optimize the parameter *θ* to minimize the discrepancy between the predicted probability 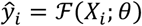 and the ground truth label *y*_*i*_. Considering that PreFoG samples are extremely sparse in natural gait (Class Imbalance), we introduce Focal Loss as the loss function *ℒ*, which forces the model to focus on hard-to-classify PreFoG samples by down-weighting the easily classified samples (normal walking). The loss function is defined as follows:

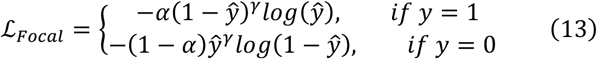

where *α* is the balancing factor, *γ* is the focusing parameter, *y* ∈ {0,1} is the ground truth label, and ŷ ∈ [0,1] is the model’s predicted probability.

At inference, the sigmoid output was interpreted as the probability of the PreFoG class and was thresholded to obtain the binary decision.

### G. Implementation and training details

To limit data leakage, we split the data at the subject level. No participant appeared in more than one of the training, validation, and test sets. Any preprocessing step that depended on the data distribution was fitted on the training set and then applied to the validation and test sets. All models were implemented under the same preprocessing protocol, subject split, and prediction horizon. Experiments were run in PyTorch with an NVIDIA RTX 5090 GPU.

Given the sparsity of FoG events during natural walking (PreFoG samples being significantly fewer than Normal samples), we adopted a dual-balancing mechanism to alleviate the class imbalance problem: during the data loading phase, we introduced a Weighted Random Sampler. By assigning a higher sampling weight *w*_1_ to the minority class (PreFoG) and a lower weight *w*_0_ to the majority class (Normal), we ensured that the ratio of positive to negative samples within each mini-batch was maintained at a dynamic equilibrium of approximately 2:8. During the optimization phase, we employed Focal Loss to replace the standard cross-entropy loss.

Models were optimized with Adam using an initial learning rate of 1e-3, a weight decay of 1e-4, and a batch size of 128. Training was stopped early when validation G-Mean no longer improved. The main hyperparameters are shown in Table I.

**TABLE I.** Summary of Implementation Details and Hyperparameters.

| Hyperparameters | Value |
| --- | --- |
| Optimizer | Adam |
| Initial Learning Rate | $1e-3$ |
| Weight Decay | $1e-4$ |
| Batch Size | 128 |
| Maximum Epochs | 100 |
| Loss Function | Focal Loss ($\gamma = 2.0$, $\alpha = 0.8$) |
| Stem Layer | Conv1d ($k=7$, $s=1$, $p=3$), BN, ReLU→64 channels |
| Inception Block 1 | Micro ($k=1$, 3), Meso ($k=1$, 5), Macro ($k=1$, 11), Pool ($k=3$), Stride = 1→128 channels |
| Inception Block 2 | Micro ($k=1$, 3), Meso ($k=1$, 5), Macro ($k=1$, 11), Pool ($k=3$), Stride = 2→128 channels |
| Bi-Mamba Encoder | $d_{model} = 128$, $d_{state} = 16$, $d_{conv} = 4$, expand=2, Output dimension = 256 |
| Skip Connection | Global Average Pooling (GAP)→128 |
| Feature Projection | Linear mapping→ $Embed_{dim} = 64$ |
| Multi-Head Attention | Number of Heads = 4, Sequence Length = 3 |

To comprehensively evaluate the model’s performance on the imbalanced dataset, in addition to the conventional accuracy and sensitivity/recall, we focus on the G-Mean (Geometric Mean) and F2-Score. The G-Mean 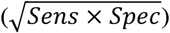 provides a balanced evaluation of the model’s recognition ability for both positive and negative classes, whereas the F2-Score assigns a higher weight to sensitivity to prioritize the minimization of clinical risks and ensure a high detection rate for freezing events. Furthermore, we report the AUC-ROC and MCC as comprehensive performance metrics.

## III. Results

### A. Statistical Analysis of Spectral Representation

We first examined whether the proposed frequency stream was supported by measurable spectral differences in the data. Preprocessed plantar pressure signals were converted to log-magnitude spectra, and three gait states were compared: normal walking, PreFoG at different lead times, and FoG. Normal walking served as the within-subject baseline. Differences across the 0-25 Hz range were tested using the Wilcoxon signed-rank test, and Bonferroni correction was applied to reduce the risk of false discoveries across frequency bins.

Fig. 4 summarizes the corrected p-values for each PreFoG stage against normal walking. The main observation is that PreFoG does not behave as a simple monotonic progression toward FoG. In the 3-8 Hz freeze-related band, the strongest departure from normal walking appears at 2-3 s before onset rather than immediately before freezing. The 0-1 s window shows a weaker contrast, suggesting that the pressure pattern at this late stage is already moving toward the freezing state and is less separable from FoG itself.

**Fig. 4.**
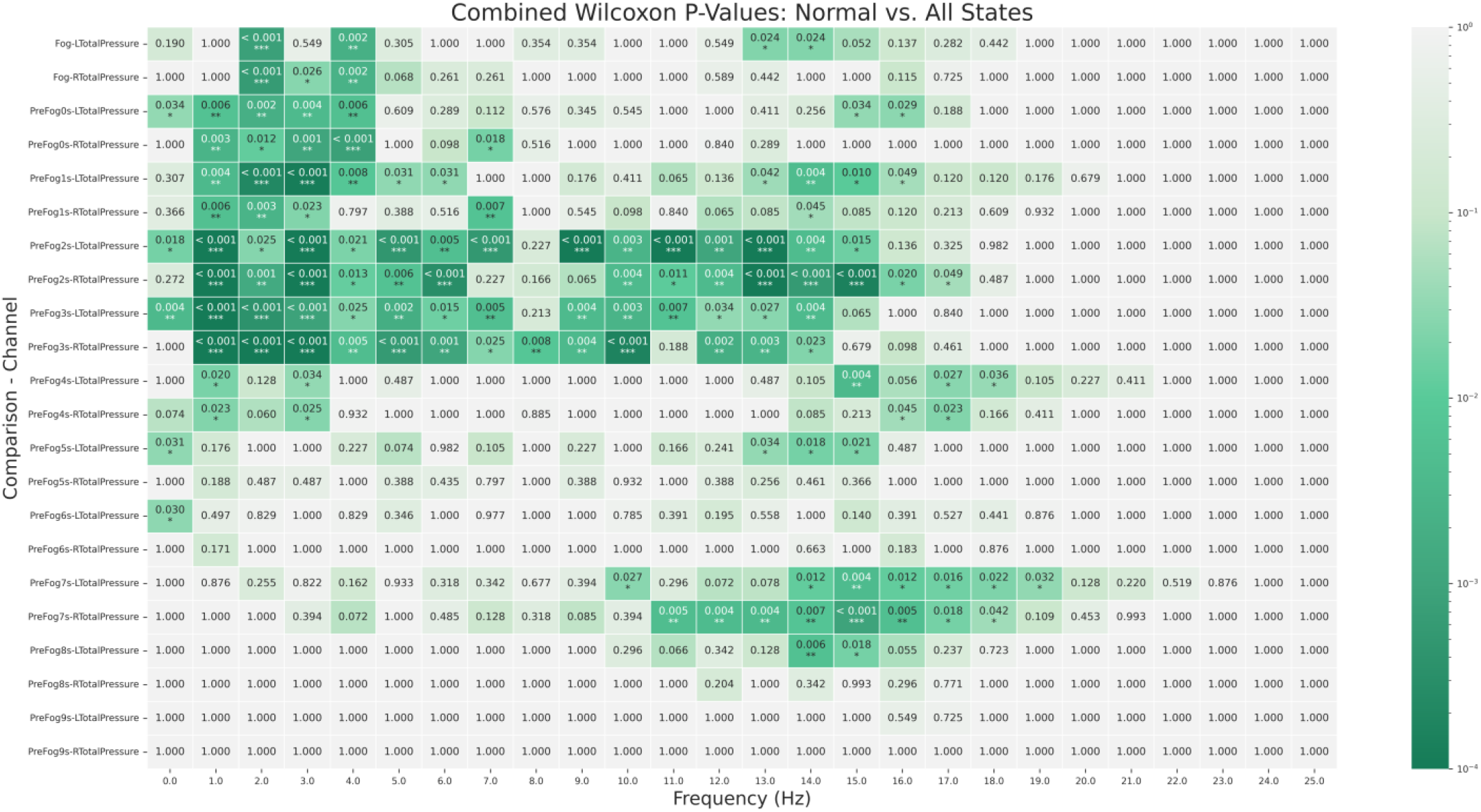
Spectrum difference heatmap (Normal vs All States)

The 8-20 Hz band also showed significant differences, especially during the 1-3 s interval. These higher-frequency components are consistent with brief impact-like or postural adjustment signals that are not present during stable walking. Localized significance was also observed around 7 s before onset in the 10-20 Hz range. Although this early effect should be interpreted cautiously, it indicates that some measurable gait perturbations may occur several seconds before the clinical FoG event.

Taken together, the heatmap supports the design of a multi-band frequency stream. The pre-freezing period contains time-varying spectral information: earlier high-frequency perturbations are followed by a stronger 3-8 Hz abnormality at 2-3 s before FoG. This pattern would likely be blurred if the full spectrum were treated as a single undifferentiated feature vector.

We then compared each PreFoG stage with the FoG stage itself to clarify how the pre-freezing signature evolves as onset approaches. Fig. 5 plots the significance curves for the left and right total plantar pressure. Rather than a gradual linear convergence, the curves show a clear non-monotonic pattern.

**Fig. 5.**
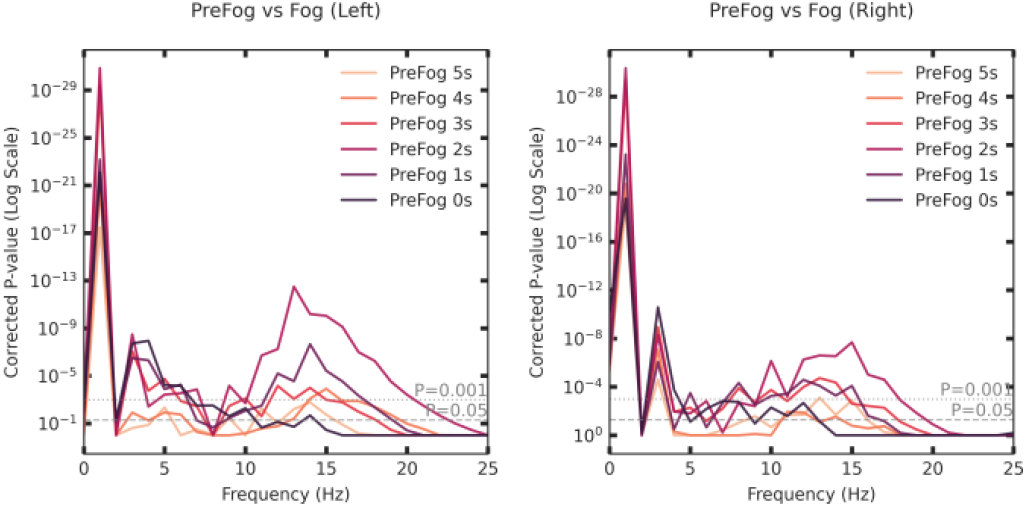
Spectrum difference heatmap (PreFog vs Fog)

At 4-5 s before onset, spectral features begin to separate from the FoG pattern. The separation becomes strongest at 2-3 s, where the corrected p-values reach their lowest values and the spectral contrast is most evident. During the final 0-1 s before onset, this contrast decreases. A plausible interpretation is that the gait signal has begun to resemble the freezing state, making the distinction between late PreFoG and FoG less pronounced than at the earlier struggle phase.

This result is important for model design and for eventual cue timing. The 2-3 s interval appears to contain a particularly informative pre-freezing signature that is neither normal walking nor fully developed FoG. A model that can attend to this interval should therefore be better suited for early warning than a model trained only to recognize the freezing state.

### B. Comparative Performance Evaluation

For the main prediction experiment, the lead time was set to 2 s and performance was evaluated on the held-out test subjects. Fig. 6 reports the confusion matrix, ROC curve, and UMAP visualization. The model correctly identified 31 of 33 PreFoG samples, corresponding to a sensitivity of 93.94%, with only two missed pre-freezing events. This high recall is important because missed FoG warnings carry a greater safety cost than occasional false alerts in a wearable assistance scenario.

**Fig. 6.**
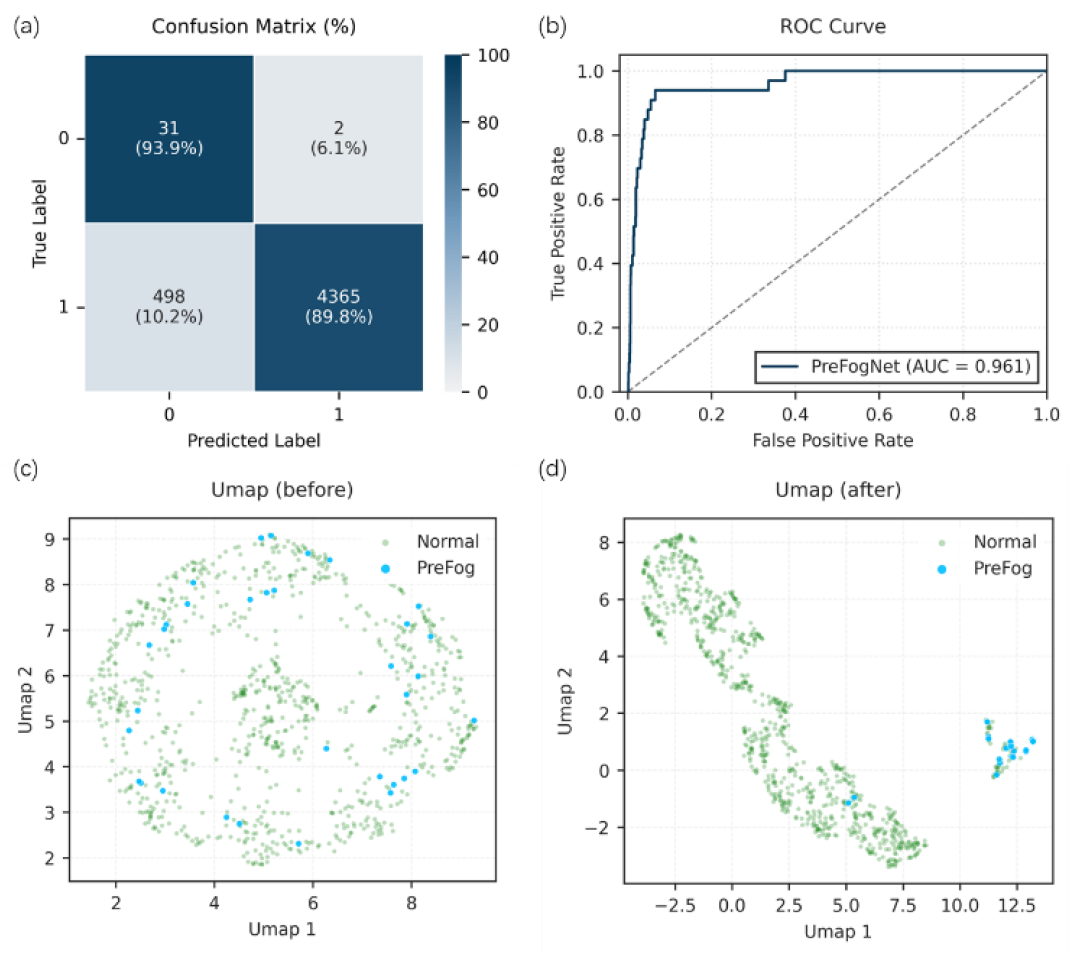
Main experimental training results

In absolute terms, the model produced 498 false-positive predictions among a much larger number of normal samples. This trade-off should not be overstated as clinically sufficient without prospective testing, but it is encouraging for an early-warning system where sensitivity is prioritized and false alarms must remain manageable. The G-Mean of 0.9183 and AUC-ROC of 0.9607 indicate that the model did not obtain high accuracy merely by following the majority class.

The UMAP plots provide a qualitative check on what the network learned. Before feature learning, normal and PreFoG windows were strongly entangled. After the dual-stream representation, PreFoG samples formed more compact clusters with clearer separation from normal walking. This visualization is not a substitute for quantitative testing, but it supports the interpretation that the model extracts discriminative time-frequency structure rather than relying on a single amplitude-related artifact.

To place these results in context, we compared PreFoGNet with 11 baselines, including classical machine-learning algorithms and deep learning models. All methods used the same subject-wise partitioning, preprocessing protocol, and 2 s prediction horizon. Tables II and III report the full results.

**TABLE II.** Compared to traditional machine learning methods.

| Methods | Acc | Sens | Spec | G-mean | F2-Score | Roc |
| --- | --- | --- | --- | --- | --- | --- |
| LR | 0.5319 | 0.7879 | 0.5301 | 0.6463 | 0.6722 | 0.7069 |
| RF | 0.7416 | 0.6667 | 0.7421 | 0.7034 | 0.6094 | 0.775 |
| SVM | 0.8056 | 0.6364 | 0.8067 | 0.7165 | 0.5947 | 0.785 |
| XGB | 0.7194 | 0.6667 | 0.7197 | 0.6927 | 0.6049 | 0.7592 |
| **Ours** | **0.8979** | **0.9394** | **0.8976** | **0.9183** | **0.9055** | **0.9607** |

**TABLE III.** Compared to mainstream deep learning methods.

| Methods | Acc | Sens | Spec | G-mean | F2-Score | Roc |
| --- | --- | --- | --- | --- | --- | --- |
| 1D-Convnet | 0.7145 | 0.6667 | 0.7148 | 0.6903 | 0.6039 | 0.7847 |
| LSTM | 0.8762 | 0.8182 | 0.8766 | 0.8469 | 0.783 | 0.8873 |
| Transformer | 0.7469 | 0.7273 | 0.7471 | 0.7371 | 0.6658 | 0.8194 |
| Uni-LSTM | 0.5596 | 0.7576 | 0.5583 | 0.6503 | 0.6521 | 0.6562 |
| CNN | 0.8433 | 0.7879 | 0.8437 | 0.8153 | 0.7455 | 0.8685 |
| LSTM+16 | 0.7349 | 0.6364 | 0.7356 | 0.6842 | 0.5805 | 0.6711 |
| TCNN | 0.8936 | 0.8182 | 0.8941 | 0.8553 | 0.7879 | 0.9495 |
| **Ours** | **0.8979** | **0.9394** | **0.8976** | **0.9183** | **0.9055** | **0.9607** |

Classical machine-learning models were less suitable for the raw multi-channel pressure sequence. SVM preserved moderate specificity but missed many PreFoG samples, whereas logistic regression achieved higher sensitivity only at the cost of very low specificity. This pattern is consistent with the nonlinear and time-varying nature of pre-freezing gait: hand-crafted or shallow decision boundaries do not easily capture the interaction between bilateral loading, local contact transients, and spectral changes.

Among the deep learning baselines, LSTM and Transformer models improved over most classical methods but remained below PreFoGNet. Relative to LSTM and 1D-Transformer, PreFoGNet increased G-Mean by 7.14% and 18.12%, respectively. The result suggests that the bidirectional Mamba component is useful for preserving longer-range pressure dynamics, while avoiding some of the optimization and data-efficiency issues that can affect recurrent or attention-based models in rare-event settings.

Overall, the comparative experiments indicate that the gain of PreFoGNet is not simply due to using a deeper network. Its advantage comes from combining complementary assumptions: local multi-scale contact encoding, long-range temporal modeling, and physiologically constrained spectral analysis.

### C. Impact of Prediction Horizon and Window Size

We next examined how performance changed with prediction horizon and input window length. Fig. 7 shows that performance remained most stable within 0-3 s before onset. In this interval, the 3 s input window kept the G-Mean above 0.91, which agrees with the spectral finding that the 2-3 s stage contains the strongest pre-freezing signature. Beyond approximately 4 s, performance gradually declined but remained informative, with G-Mean values above 0.88.

**Fig. 7.**
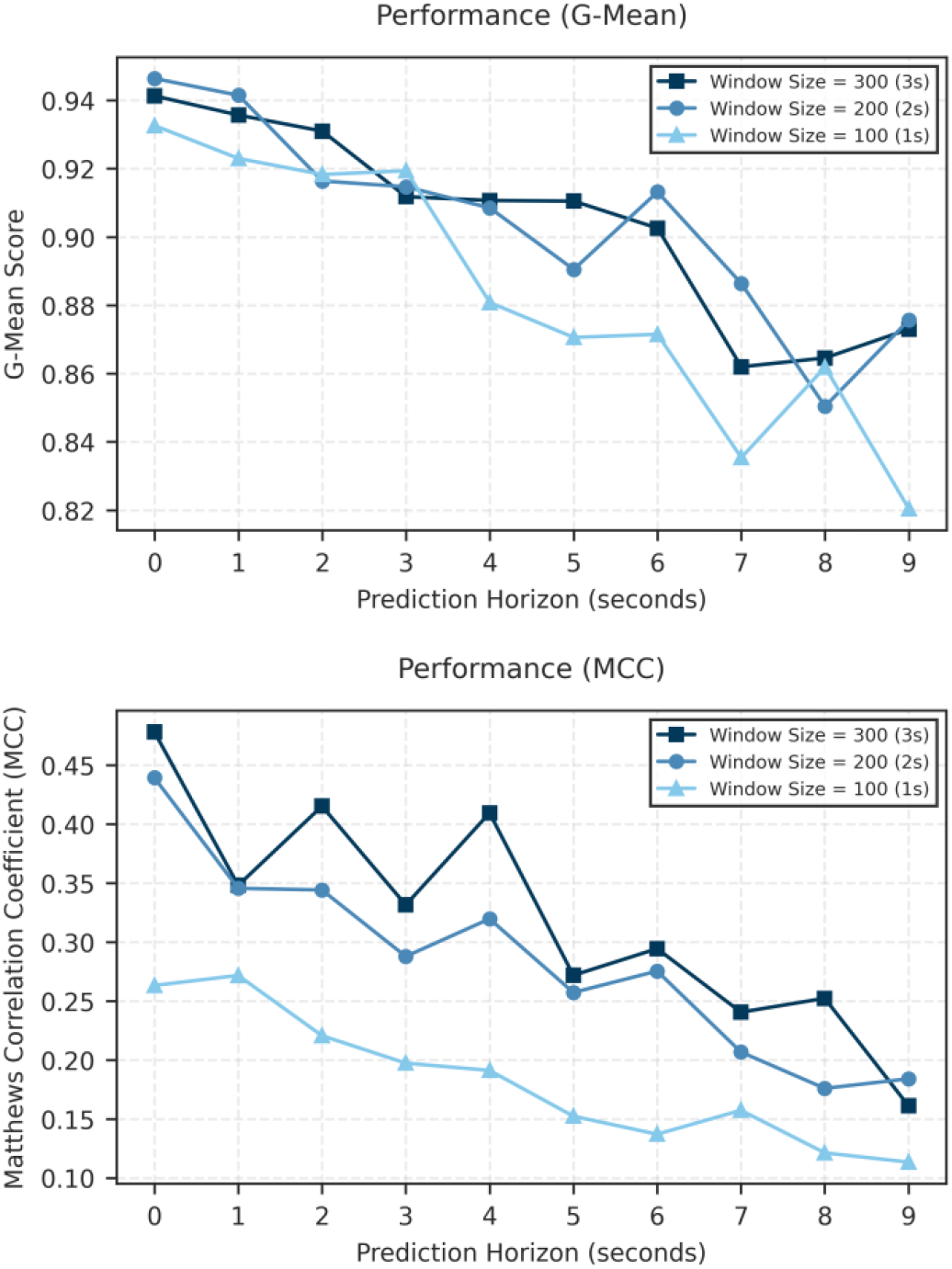
Model performance metrics under different lead times and window sizes

Input length also mattered. The 3 s context generally outperformed the 1 s and 2 s contexts, especially for MCC, which is sensitive to imbalanced classification. This indicates that a longer pressure history helps the model integrate several gait cycles and distinguish progressive deterioration from isolated contact irregularities. The shorter 1 s window has lower latency and computational cost, but it appears to truncate useful prodromal information.

### D. Model Robustness Against Noise Interference

Wearable pressure sensors may be affected by electronic noise, shoe movement, loose fitting, and slow baseline shifts. To test whether the model was overly dependent on clean laboratory signals (*T*_*horizon*_ *=* 2*s, L =* 100), we added Gaussian noise, uniform noise, and baseline drift at increasing intensities. Gaussian and uniform noise mainly disturbed the high-frequency spectrum, whereas baseline drift changed the temporal offset with relatively little spectral distortion (Fig. 8).

**Fig. 8.**
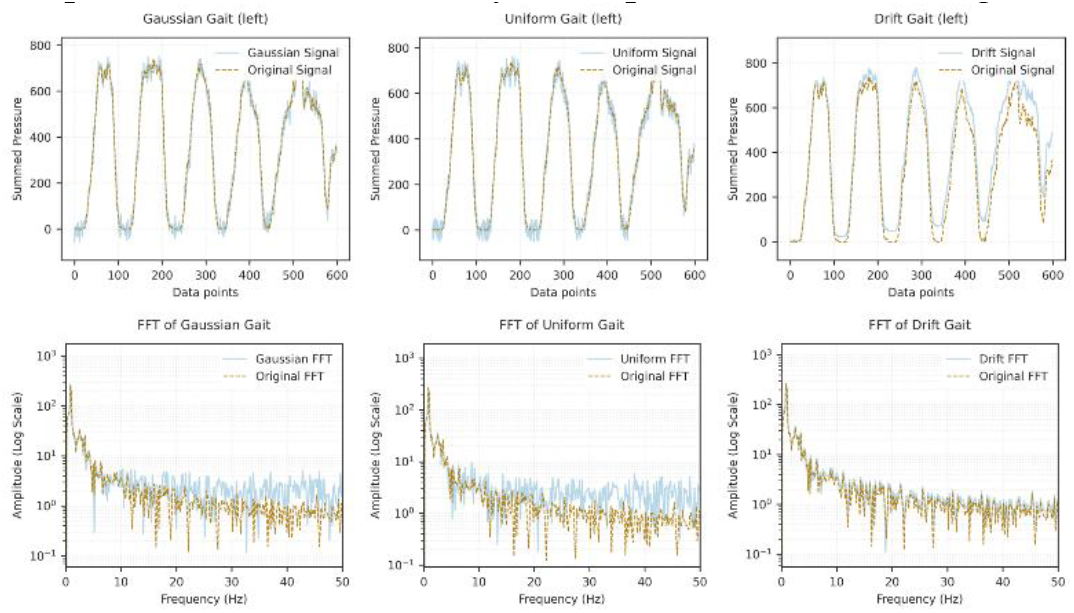
The impact of noise on the original signal

As shown in Fig. 9, low-intensity noise (σ *=* 0.01) produced only a small decrease in G-Mean relative to the clean baseline of 0.9183. At medium intensity (σ *=* 0.05), the model retained more than 85% of its baseline performance across all three noise types.At higher noise levels (σ *=* 0.1), the model was more sensitive to Gaussian noise than to uniform noise or drift. This is reasonable because Gaussian noise can introduce larger transient spikes, which may mimic brief abnormal contacts or mask the weak 3-8 Hz pre-freezing components. Baseline drift was less harmful, probably because the spectral stream remained relatively stable and the temporal stream could use longer context to absorb slow offsets.

**Fig. 9.**
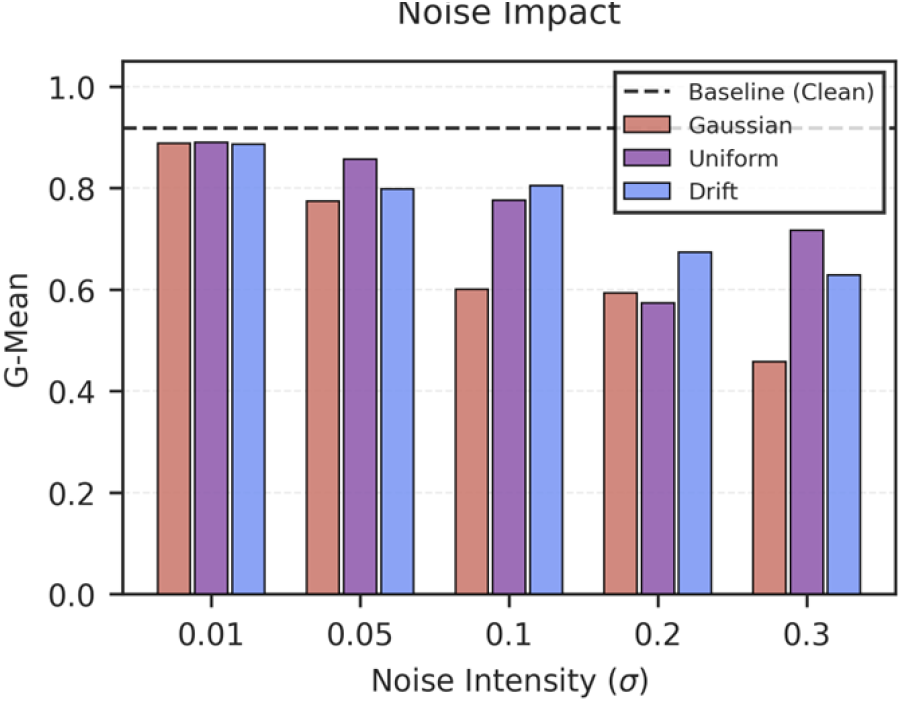
The Influence of Different Intensity Noise on Model Performance

These experiments do not replace validation with real-world sensor artifacts, but they indicate that the model has some tolerance to common signal degradations expected in wearable plantar-pressure systems.

### E. Fault tolerance to single-channel loss

We also tested the effect of losing a single pressure channel, a realistic failure mode during long-term insole use. Each channel was removed and reconstructed using the mean of its physically adjacent sensors, allowing us to estimate both fault tolerance and regional importance. Fig. 10 shows the percentage drop in G-Mean after each channel was removed and imputed. Larger drops indicate channels whose information cannot be easily recovered from neighboring sensors, whereas small or negative drops indicate local redundancy.

**Fig. 10.**
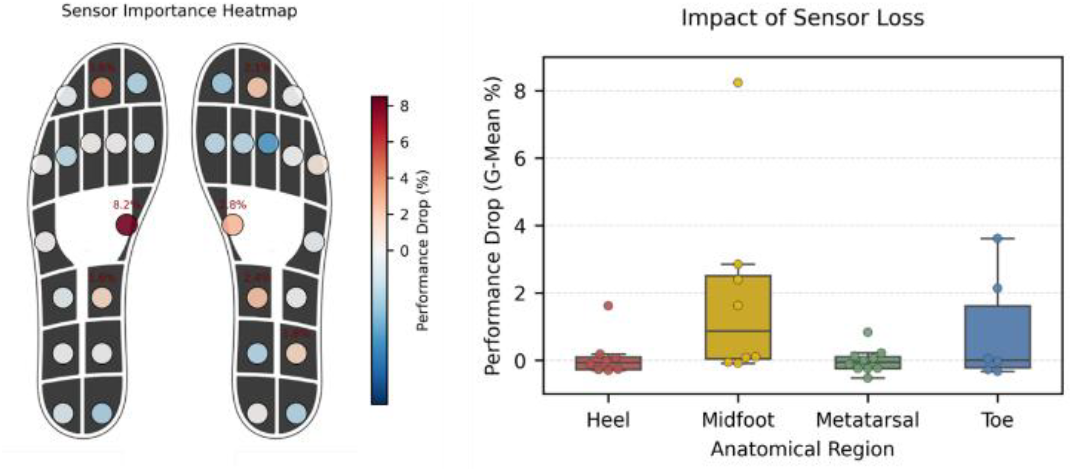
The impact of channel loss on model performance

The largest drops occurred in the midfoot region, especially Ch 7 on the left foot and Ch 23 on the right foot, where G-Mean decreased by 8.24% and 2.85%, respectively. This is biomechanically plausible. The plantar arch does not behave like a simple average of the heel and forefoot; it reflects load transfer and balance regulation during stance.

Toe and lateral forefoot sensors also contributed to prediction. The paired importance of Ch 15 and Ch 31 suggests that the model uses bilateral information rather than relying on one foot. This fits the clinical observation that FoG often involves disturbed weight shifting and impaired step initiation, processes that are expressed through both toe-off dynamics and inter-limb loading.

A few channels showed a slight improvement after imputation. We interpret this as a smoothing effect rather than true evidence that these sensors are unnecessary. When a less informative channel is replaced by the average of nearby informative channels, local noise may be reduced, leading to a small performance gain. For deployment, the result still argues for preserving sensor quality in the midfoot and toe regions.

### F. Component performance evaluation

The ablation study was designed to determine which parts of PreFoGNet were responsible for the performance gain. As shown in Table IV, removing the frequency stream caused the largest reduction in G-Mean, showing that spectral information is central to separating PreFoG from negative examples. Using a raw FFT representation partially recovered performance, but remained below the proposed band-wise spectral module, indicating that physiologically organized bands provide more useful structure than a global spectrum alone.

**TABLE IV.** Compared to traditional machine learning method.

| Exp. | Inception | BiMamba | Spectrum | Acc | Sens | Spec | G-Mean | ΔG-Mean |
| --- | --- | --- | --- | --- | --- | --- | --- | --- |
| Exp.1 | ✓ | ✓ | × | 0.8231 | 0.9394 | 0.8223 | 0.8789 | ↓ 4.29% |
| Exp.2 | ✓ | × | ✓ | 0.8311 | 0.9394 | 0.8304 | 0.8832 | ↓ 3.82% |
| Exp.3 | ✓ | Bi-LSTM | ✓ | 0.8417 | 0.9394 | 0.841 | 0.8889 | ↓ 3.20% |
| Exp.4 | × | ✓ | ✓ | 0.857 | 0.9091 | 0.8567 | 0.8825 | ↓ 3.90% |
| Exp.5 | ✓ | ✓ | FFT | 0.8638 | 0.9394 | 0.8633 | 0.9005 | ↓ 1.94% |
| **Ours** | ✓ | ✓ | ✓ | **0.8979** | **0.9394** | **0.8976** | **0.9183** | - |

The temporal module also contributed substantially. Removing Bi-Mamba reduced G-Mean by 3.82%, and replacing it with Bi-LSTM reduced G-Mean by 3.20%. These results support the use of a state-space temporal model for plantar pressure sequences, where the relevant changes may develop gradually and where computational efficiency is important for later wearable implementation.

Finally, replacing the multi-scale Inception block with a single-scale convolution reduced sensitivity from 93.94% to 90.91%. This suggests that the model benefits from viewing the signal at multiple temporal scales: short kernels capture impact-like transients, whereas longer receptive fields capture gait-cycle-level changes.

## IV. Discussion

This work proposes PreFoGNet, a dual time-frequency framework for early FoG prediction from bilateral plantar pressure. The main finding is that plantar pressure contains a usable pre-freezing signature when modeled jointly in the temporal and spectral domains. Under subject-wise testing at a 2 s lead time, PreFoGNet achieved 93.94% sensitivity, 89.76% specificity, a G-Mean of 0.9183, and an AUC-ROC of 0.9607, outperforming classical and deep learning baselines.

The temporal stream addresses a practical limitation of many sequence models used for FoG prediction. The multi-scale Inception encoder captures local contact morphology at different durations, while Bi-Mamba provides a compact mechanism for modeling longer pressure histories. The ablation results show that both parts are needed: removing the multi-scale encoder mainly reduced sensitivity, whereas removing or replacing Bi-Mamba reduced balanced performance. This supports the interpretation that PreFoG prediction requires both local waveform detail and longer-term gait context.

The frequency-stream results provide a second explanation for the model’s behavior. The spectral analysis showed that the 3-8 Hz band was most distinctive at 2-3 s before onset and became less separable during the last second before FoG. This pattern is consistent with a transition in which patients first show compensatory, high-frequency stepping or trembling-like activity and then move closer to motor arrest. By separating locomotion, freeze-related, and high-frequency bands, the model can use these changes without forcing the network to discover the entire physiological structure from data alone.

The horizon analysis has implications for cue timing. The most reliable window was 0-3 s before onset, especially around 2-3 s, which may be suitable for immediate rhythmic cueing. The model still retained information at 4-6 s, but performance became less stable beyond 7 s. We therefore view 6-7 s as an approximate upper boundary for plantar-pressure-only prediction in this dataset, not as a guaranteed warning time for every patient.

Robustness experiments further support the feasibility of the signal-processing approach. The model tolerated moderate Gaussian noise, uniform noise, baseline drift, and single-channel loss, suggesting that it did not rely on a single fragile feature. The channel-loss analysis also gave a biomechanically meaningful result: midfoot and toe regions were most important. These areas are involved in center-of-pressure transfer and step initiation, which are known to be disrupted around FoG.

Several limitations remain. First, the experiments used data collected under controlled walking tasks. Free-living walking includes stairs, crowds, narrow spaces, and irregular footwear conditions, all of which may change the false-alarm profile. Second, although Mamba is computationally attractive, actual deployment will require latency measurement, and testing on embedded hardware. These issues should be addressed before clinical claims are made about real-time closed-loop prevention.

## Data Availability

All data produced in the present work are contained in the manuscript. The WearGait-PD dataset is available on Synapse (SAGE Bionetworks) at the following URL: https://www.synapse.org/Synapse:syn52540892/wiki/623751.

## V. Code availability

The source code of PreFoGNet is available on GitHub (https://github.com/Ivan020121/PreFoGNet).

## References

[1] A. Samii, J. G. Nutt, and B. R. Ransom, “Parkinson’s disease,” The Lancet, vol. 363, no. 9423, pp. 1783–1793, 2004.

[2] Z.-X. Zhang, G. C. Roman, Z. Hong, C.-B. Wu, Q.-M. Qu, J.-B. Huang, B. Zhou, Z.-P. Geng, J.-X. Wu, H.-B. Wen, H. Zhao, and G. E. P. Zahner, “Parkinson’s disease in China: prevalence in Beijing, Xian, and Shanghai,” The Lancet, vol. 365, no. 9459, pp. 595–597, 2005/02/12/, 2005.

[3] B. R. Bloem, J. M. Hausdorff, J. E. Visser, and N. Giladi, “Falls and freezing of gait in Parkinson’s disease: A review of two interconnected, episodic phenomena,” Movement Disorders, vol. 19, no. 8, pp. 871–884, 2004/08/01, 2004.

[4] N. Giladi, R. Kao, and S. Fahn, “Freezing phenomenon in patients with parkinsonian syndromes,” Movement Disorders, vol. 12, no. 3, pp. 302–305, 1997/05/01, 1997.

[5] S. Perez-Lloret, L. Negre-Pages, P. Damier, A. Delval, P. Derkinderen, A. Destée, W. G. Meissner, L. Schelosky, F. Tison, and O. Rascol, “Prevalence, Determinants, and Effect on Quality of Life of Freezing of Gait in Parkinson Disease,” JAMA Neurology, vol. 71, no. 7, pp. 884–890, 2014.

[6] S. Pardoel, J. Kofman, J. Nantel, and E. D. Lemaire, “Wearable-Sensor-Based Detection and Prediction of Freezing of Gait in Parkinson’s Disease: A Review,” Sensors, 19, 2019].

[7] G. Shalin, S. Pardoel, J. Nantel, E. D. Lemaire, and J. Kofman, “Prediction of Freezing of Gait in Parkinson’s Disease from Foot Plantar-Pressure Arrays using a Convolutional Neural Network.” pp. 244–247.

[8] G. Shalin, S. Pardoel, E. D. Lemaire, J. Nantel, and J. Kofman, “Prediction and detection of freezing of gait in Parkinson’s disease from plantar pressure data using long short-term memory neural-networks,” Journal of NeuroEngineering and Rehabilitation, vol. 18, no. 1, pp. 167, 2021/11/27, 2021.

[9] S. Pardoel, A. AlAkhras, E. Jafari, J. Kofman, E. D. Lemaire, and J. Nantel, “Real-Time Freezing of Gait Prediction and Detection in Parkinson’s Disease,” Sensors, vol. 24, no. 24, pp. 8211, 2024.

[10] M. Mancini, B. R. Bloem, F. B. Horak, S. J. G. Lewis, A. Nieuwboer, and J. Nonnekes, “Clinical and methodological challenges for assessing freezing of gait: Future perspectives,” Movement Disorders, vol. 34, no. 6, pp. 783–790, 2019/06/01, 2019.

[11] N. Yanagisawa, R. Hayashi, and H. Mitoma, “Pathophysiology of frozen gait in Parkinsonism,” Advances in neurology, vol. 87, pp. 199–207, 2001, 2001.

[12] S. Pardoel, J. Nantel, J. Kofman, and E. D. Lemaire, “Prediction of Freezing of Gait in Parkinson’s Disease Using Unilateral and Bilateral Plantar-Pressure Data,” Frontiers in Neurology, vol. Volume 13 – 2022, 2022-April-28, 2022.

[13] C. Cosentino, M. Putzolu, S. Mezzarobba, M. Cecchella, T. Innocenti, G. Bonassi, A. Botta, G. Lagravinese, L. Avanzino, and E. Pelosin, “One cue does not fit all: A systematic review with meta-analysis of the effectiveness of cueing on freezing of gait in Parkinson’s disease,” Neuroscience & Biobehavioral Reviews, vol. 150, pp. 105189, 2023/07/01/, 2023.

[14] A. J. Anderson, D. Eguren, M. A. Gonzalez, M. Caiola, N. Khan, S. Watkinson, I. Zuccaroli, S. S. Hirczy, C. P. Zabetian, K. Mills, E. Moukheiber, L. Moro-Velazquez, N. Dehak, C. Motley, B. C. Muir, A. Butala, and K. Kontson, “WearGait-PD: An Open-Access Wearables Dataset for Gait in Parkinson’s Disease and Age-Matched Controls,” Scientific Data, vol. 13, no. 1, pp. 440, 2026/02/12, 2026.

[15] H. Ismail Fawaz, B. Lucas, G. Forestier, C. Pelletier, D. F. Schmidt, J. Weber, G. I. Webb, L. Idoumghar, P.-A. Muller, and F. Petitjean, “InceptionTime: Finding AlexNet for time series classification,” Data Mining and Knowledge Discovery, vol. 34, no. 6, pp. 1936–1962, 2020/11/01, 2020.

[16] A. Gu, and T. Dao, “Mamba: Linear-Time Sequence Modeling with Selective State Spaces,” in Conference on Language Modeling, 2024.

[17] A. Nieuwboer, R. Dom, W. De Weerdt, K. Desloovere, S. Fieuws, and E. Broens-Kaucsik, “Abnormalities of the spatiotemporal characteristics of gait at the onset of freezing in Parkinson’s disease,” Movement Disorders, vol. 16, no. 6, pp. 1066–1075, 2001/11/01, 2001.

[18] A. Vaswani, N. Shazeer, N. Parmar, J. Uszkoreit, L. Jones, A. N. Gomez, Ł. Kaiser, and I. Polosukhin, “Attention is all you need,” in Proceedings of the 31st International Conference on Neural Information Processing Systems, Long Beach, California, USA, 2017, pp. 6000–6010.

